# Knowledge of antimicrobial Resistance in Bangladeshi Caregivers of School-going Children: A Cross-sectional study

**DOI:** 10.64898/2026.11.21.26368630

**Authors:** Ruhi Tamanna Chowdhury, Umme Habiba Asamoni, Rodayla Raiyan, Sayeda Hafsa, Shahed Anan Sajeeb, Ubarna Bhattacharjee

## Abstract

**Background:** Antimicrobial resistance (AMR) is a critical global health issue exacerbated by inappropriate antibiotic use. This study aimed to assess AMR awareness and antibiotic practices among caregivers in Bangladesh.

**Methods:** A cross-sectional design was employed with structured questionnaires administered to 225 caregivers. The survey evaluated participants’ knowledge of AMR, antibiotic usage behaviors, information sources, and socio-demographic variables.

**Results:** Results demonstrated a poor understanding of AMR: 78% recognized antibiotics as effective against bacterial infections, while only 35% acknowledged their ineffectiveness against viruses. In terms of antibiotic practices, 30% discontinued treatment prematurely, and 40% engaged in self-medication, primarily influenced by pharmacy advice (45%) or previous prescriptions (30%). Healthcare professionals were the predominant source of information (55%), followed by media (30%) and community programs (15%). Knowledge disparities were observed among socio-demographic groups, with higher awareness in educated, urban, and affluent caregivers (p<0.05).

**Conclusions:** The study highlights significant gaps between AMR knowledge and practices, particularly regarding antibiotic misuse and viral infections. It underscores the urgent need for targeted educational efforts, enhanced prescription regulation, and improved healthcare access to mitigate AMR risks in Bangladesh. Behavioral interventions addressing knowledge deficiencies and practical challenges are essential to combat this growing public health issue.

## 1. Introduction

Antimicrobial resistance (AMR) emerges when microorganisms, including bacteria, viruses, fungi, and parasites, develop the ability to withstand the effects of antimicrobial drugs, thereby complicating infection treatment (Iwu & Patrick, 2021). This resistance arises from genetic mutations or the acquisition of resistance genes, diminishing the effectiveness of previously successful medications (Laxminarayan et al., 2013). The swift escalation of AMR has been recognized as a significant global public health issue, threatening modern medicine and elevating the risk of severe infections, prolonged illness, and increased mortality rates (Singer et al., 2016). According to the Global Burden of Disease (GBD) study, AMR was primarily responsible for an estimated 1.27 million deaths worldwide in 2019, with middle- and low-income countries experiencing the most severe impact (Murray et al., 2022). Without prompt intervention, AMR could result in 10 million deaths annually by 2050, surpassing cancer as the leading cause of death (Singer et al., 2016).

Antimicrobial resistance (AMR) has profound implications that extend beyond public health, adversely affecting economic stability and global healthcare systems. A particularly alarming consequence is the ineffectiveness of conventional therapies or standard treatments, leading to prolonged infections, increased severity of illnesses, and heightened risks of complications (Lee Ventola, 2015). Common infections, such as tuberculosis (TB), pneumonia, and urinary tract infections, are increasingly unresponsive to first-line antibiotics, resulting in extended hospitalizations and elevated mortality rates (Iwu & Patrick, 2021). Moreover, AMR escalates healthcare costs by necessitating the use of more expensive and toxic alternative therapies (Dadgostar, 2019). Research has projected that infections related to AMR could increase global healthcare expenditures by 100 trillion dollars by 2050, impacting both low- and high-income countries (Singer et al., 2016). Additionally, critical medical procedures, including surgeries, organ transplants, and chemotherapy, which rely on effective antibiotics to prevent infections, are jeopardized by the rise of drug-resistant pathogens (Cassini et al., 2019). The primary drivers of AMR are human behaviors, notably the misuse and overuse of antibiotics in both medical and agricultural sectors (Holmes et al., 2016). Resistance proliferates more rapidly when antibiotics are misused, such as being taken without prescriptions, used for viral infections like the flu or common cold, or when prescribed courses are not completed (Llor & Bjerrum, 2014). In many countries, antibiotics are available over the counter without adequate regulation, leading to widespread misuse, particularly in low-income regions (Founou et al., 2016). Furthermore, the misuse of antibiotics in agriculture and livestock exacerbates AMR by promoting the dissemination of resistant bacteria through the environment and food supply (Van Boeckel et al., 2015). Altering public perceptions and increasing awareness about responsible antibiotic use is a significant challenge in addressing AMR, as behavioral interventions have shown promising results in reducing the use of unnecessary antibiotics.

Caregivers play a pivotal role in the healthcare decision-making process for children, particularly concerning antibiotic usage (World Health Organization, 2021). Their perceptions and knowledge significantly influence the prudent use of antibiotics, which is essential in addressing antimicrobial resistance (AMR). Understanding the appropriate use of antibiotics can deter misuse, over-prescription, and self-medication, thereby reducing the risk of antibiotic resistance. Many caregivers erroneously believe that antibiotics are necessary for all illnesses, including viral infections such as influenza and the common cold (Centers for Disease Control and Prevention, 2020). This misconception leads to antibiotic overuse, diminishing their effectiveness against bacterial infections and promoting resistance. Educating caregivers about the correct indications for antibiotic use is a crucial strategy to address this issue. In Bangladesh, the problem is exacerbated by the over-the-counter availability of antibiotics (World Health Organization, 2021). For reasons of convenience, lack of knowledge, or financial considerations, caregivers often resort to self-medication. This practice frequently results in inappropriate antibiotic selection, incomplete courses, and incorrect dosages, all of which contribute to the escalating problem of AMR. To mitigate these risks, there is a need for stricter legislation regulating antibiotic sales and increased public awareness. Additionally, caregivers’ education, socioeconomic status, and access to healthcare significantly impact their antibiotic-related decisions. Individuals with limited healthcare access may resort to self-medication, whereas caregivers with higher education levels are more likely to recognize the risks associated with antibiotic overuse. AMR can be reduced by empowering caregivers to make informed decisions through initiatives aimed at raising awareness, improving healthcare access, and providing appropriate education. In conclusion, caregiver education is vital in preventing antibiotic misuse and combating AMR. To promote the judicious use of antibiotics and safeguard public health, misconceptions must be dispelled, antibiotic supply regulated, and access to healthcare and education ensured.

Antimicrobial resistance (AMR) poses a significant threat to global health, particularly in low- and middle-income countries such as Bangladesh. According to research conducted by the World Health Organization (WHO) in 2019, AMR accounts for approximately 700,000 deaths annually. By 2030, it is projected to contribute to an estimated 10 million deaths worldwide (Roope et al., 2019). Various studies on antimicrobial resistance have demonstrated a steady increase in both antibiotic usage and AMR. The WHO has expressed concern that AMR may herald a post-antimicrobial era in which routine diseases could become fatal (Islam et al., 2024). The prevalence of self-medication and frequent antibiotic prescriptions for common childhood illnesses, such as colds and respiratory infections, among the Bangladeshi population is closely linked to this rise in antimicrobial resistance. Evidence indicates that uncontrolled access to antibiotics, with 18% of households in Dhaka obtaining them without a prescription, promotes widespread self-medication for mild illnesses, accelerates the emergence of AMR, and poses a significant risk to both individuals and society (Chowdhury et al., 2009). Several factors have been suggested to contribute to the inappropriate use of antibiotics in children, including excessive prescribing by physicians, parental lack of knowledge regarding proper usage, and the ease of access to antibiotics for self-medication (Wang et al., 2019). Public awareness initiatives regarding AMR and prudent antibiotic use often fail to provide comprehensive information, resulting in only a superficial understanding of the issue. These initiatives also frequently overlook cultural norms related to antibiotics and neglect vulnerable populations, such as those with limited health literacy. Healthcare professionals play a crucial role in promoting prudent use, yet they often do not engage in these initiatives. Moreover, advertisements typically do not persist long enough to effect long-term behavioral change. This study primarily focuses on the knowledge of antibiotic resistance among caregivers of children and the urgent need for effective interventions, examining the impact of AMR on children’s health in Bangladesh.

Antimicrobial resistance (AMR) is a growing public health crisis in Bangladesh, with approximately 70% of Intensive Care Unit (ICU) deaths linked to AMR-related infections (The Daily Star, 2023). The widespread misuse and overuse of antibiotics has significantly contributed to this crisis. While healthcare providers and policymakers are increasingly focusing on AMR, a critical gap remains in understanding how caregivers, especially parents of school-going children, perceive and use antibiotics. The knowledge and awareness of caregivers play a crucial role in determining antibiotic consumption patterns in children. A study in rural Bangladesh found that unqualified medical practitioners and pharmacy shopkeepers often had inadequate knowledge of antibiotic use, leading to inappropriate prescribing and dispensing practices (Haque et al., 2023). This indicates that caregivers who frequently rely on informal sources for medical advice may unintentionally contribute to the misuse of antibiotics. Therefore, assessing caregivers’ knowledge is essential for identifying misconceptions and behavioral patterns that exacerbate AMR. The findings from such assessments can inform the development of targeted public health interventions. Tailored awareness programs can help to address specific knowledge gaps and promote responsible antibiotic use among caregivers. Community engagement initiatives in Bangladesh have shown the potential to enhance AMR-related knowledge and encourage proper antibiotic usage (Chowdhury et al., 2024). By focusing on caregivers of school-going children, interventions can be more precise and effective, reducing antibiotic misuse in this vulnerable population, and ultimately mitigating the progression of AMR.

The main aim of this study was to evaluate the knowledge, awareness, and perceptions of antimicrobial resistance (AMR) among Bangladeshi caregivers of school-going children, using a cross-sectional approach. This study has several specific objectives. To evaluate caregivers’ understanding of AMR, including its causes, consequences, and impact on public health. To investigate caregivers’ antibiotic use practices, such as adherence to prescriptions and self-medication tendencies. To identify the sources from which caregivers acquire information about AMR, such as healthcare professionals, media, and community awareness initiatives. To examine socio-demographic factors influencing caregivers’ knowledge of AMR, such as education, income, geographic location, and access to healthcare

## 2. Method

### 2.1 Study Design

To address our research questions, we conducted a cross-sectional survey of antimicrobial resistance (AMR) among Bangladeshi caregivers of school-going children from Dhaka districts of Bangladesh between 15 January 2025 and 10 April 2025.

### 2.2 Target Population and Sampling

Approximately 225 caregivers of school-going children, aged 18–32 years, participated. Data collection took place at selected primary and secondary schools in Dhaka city.

The study sites (primary or secondary schools) and caregivers were selected based on the purposive sampling method. After that, the caregivers were selected randomly and approached to determine if they were available and interested in participating in the study. Once they agreed to participate voluntarily, we collected data from them using a face-to-face questionnaire. Those who were not interested, did not have sufficient time, or did not agree to provide written consent were excluded from the study.

### 2.3 Measures

A standardized questionnaire that included questions on the participants’ sociodemographic traits as well as several questions to evaluate their knowledge, attitudes, and practices regarding the use of antibiotics and the functioning of antimicrobial resistance (AMR) was used for the study. The questionnaire was created by Azim (2023) based on a detailed review of literature from various settings, with some adjustments made to ensure its relevance to the Bangladeshi context. The questionnaire comprised four primary sections:

1. Sociodemographic details about the participants, including age, gender, residence, education, occupation, monthly income, and monthly expenditures.
2. Eight questions regarding their knowledge about the use of antibiotics and AMR.
3. Eight questions regarding their attitudes towards the use of antibiotics and AMR.
4. Twelve questions about self-reported practices related to antibiotic use.

To assess their knowledge, participants were asked to respond with “Yes,” “No,” or “Do not know.” Attitudes were assessed using “Agree,” “Neutral,” or “Disagree” options. Practices related to antibiotic use were assessed through simple nominal or categorical (Yes or No) responses.

### 2.4 Statistical/Data Analysis

Data analysis for this study was performed using SPSS version 29.0 software. Descriptive statistics were first calculated to summarize sample demographics and overall scores on the original and translated Antimicrobial Resistance Scale (AMR). Pearson correlation coefficients were then calculated to assess the relationship between items in the original and translated versions, providing information about the consistency of responses across both scales.

### 2.5 Ethical Consideration

The study was conducted in compliance with ethical standards and received approval from the Institutional Review Board (IRB) at the Bangladesh Institute of Innovative Health Research. Adherence to the Declaration of Helsinki was ensured throughout the research process.

Permissions were obtained from the participants, and informed consent was secured from caregivers. Participants were assured of the confidentiality and anonymity of their responses and were informed of their right to withdraw from the study at any time without facing any adverse consequences.

## 3. Result

### 3.1 Descriptive Statistics

A total of 225 individuals were surveyed. The average age was 39.5 years (SD = 11.2). The gender composition consisted of 52% females and 48% males. Regarding educational attainment, 45% had completed secondary education, while 22% were illiterate. A significant majority (68%) lived in urban regions. The average monthly family income was □35,000 (SD = □18,000). Table 1 represents participant Demographics and Figure 01 shows Frequency distribution of different categories.

**Table 1.**
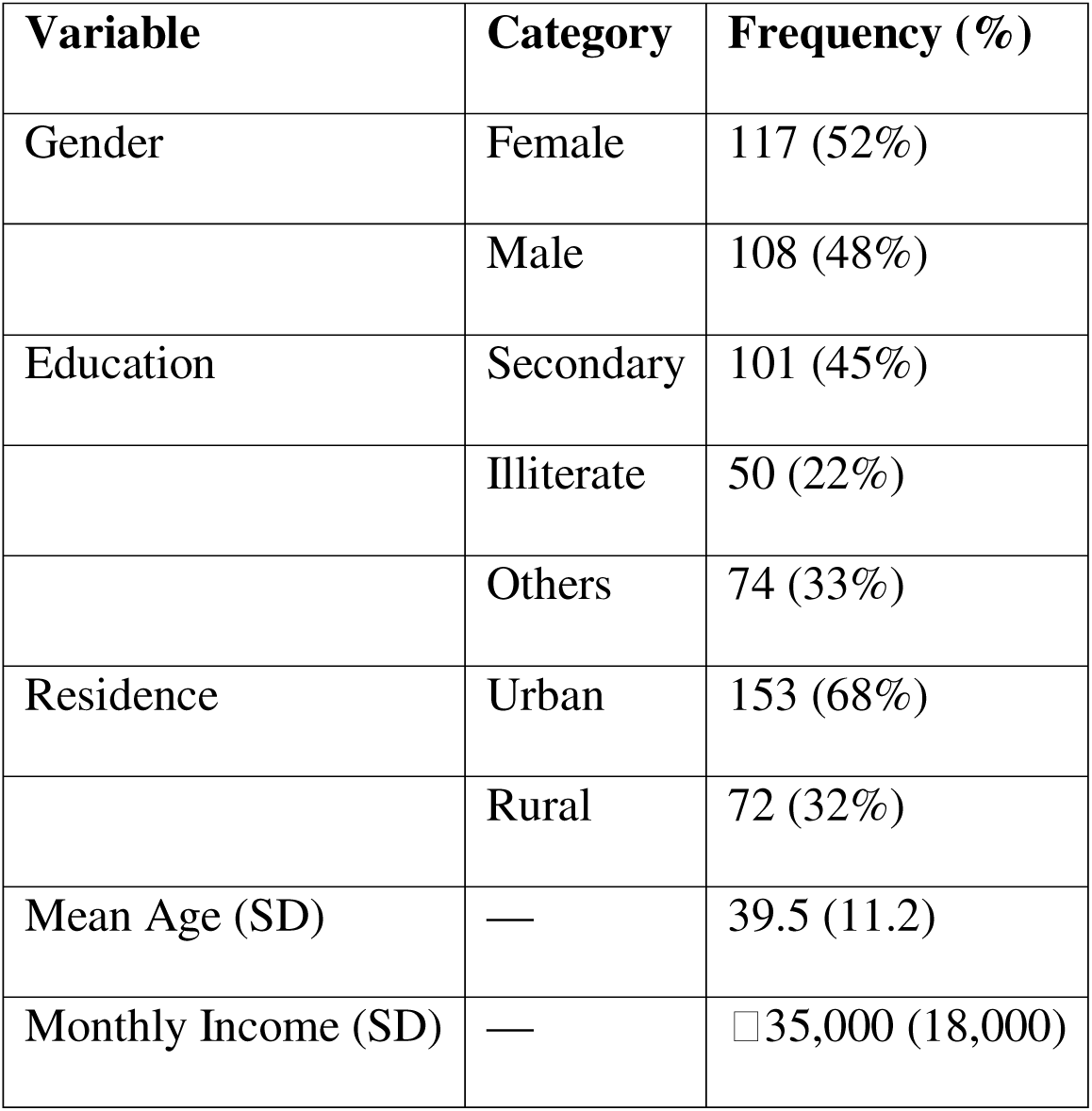
Participant Demographics.

| **Variable** | **Category** | **Frequency (%)** |
|---|---|---|
| Gender | Female | 117 (52%) |
|  | Male | 108 (48%) |
| Education | Secondary | 101 (45%) |
|  | Illiterate | 50 (22%) |
|  | Others | 74 (33%) |
| Residence | Urban | 153 (68%) |
|  | Rural | 72 (32%) |
| Mean Age (SD) | — | 39.5 (11.2) |
| Monthly Income (SD) | — | □35,000 (18,000) |

### 3.2 Knowledge of Antimicrobial Resistance (AMR)

Participants displayed diverse levels of knowledge regarding antimicrobial resistance (AMR). While 78% accurately recognized antibiotics’ efficacy against bacterial infections, only 35% understood their ineffectiveness against viral diseases. In terms of factors contributing to resistance, 62% acknowledged that missed doses could lead to resistance, and 41% identified overuse as a significant factor. Nevertheless, prevalent misconceptions persisted: 55% erroneously believed antibiotics facilitate recovery from colds and coughs, and 48% considered early cessation of antibiotics permissible. Table 2 represents Knowledge about AMR.

**Table 2:** Knowledge About AMR.

| **Item** | **Correct (%) / Misconception (%)** |
|---|---|
| Antibiotics effective against bacteria | 78% (Correct) |
| Antibiotics ineffective against viruses | 35% (Correct) |
| Missing doses cause resistance | 62% (Correct) |
| Overuse causes resistance | 41% (Correct) |
| Antibiotics cure colds/coughs | 55% (Misconception) |
| Early stop acceptable | 48% (Misconception) |

### 3.3 Antibiotic Use Practices

Antibiotic adherence exhibited variability. Sixty-five percent of subjects completed the prescribed regimen, whereas 30% ceased use prematurely upon symptom resolution. Additionally, 40% engaged in self-medication with antibiotics within the previous year, predominantly based on pharmacy suggestions (45%) or prior prescriptions (30%). Table 3 shows antibiotics Use Practice.

**Table 3:** Antibiotic Use Practices.

| **Practice** | **Frequency (%)** |
|---|---|
| Completed prescribed course | 65% |
| Stopped when feeling better | 30% |
| Self-medicated (past year) | 40% |
| Source: Pharmacy recommendation | 45% (of 40%) |
| Source: Old prescription | 30% (of 40%) |

### 3.4 Sources of AMR Information

Healthcare professionals comprised the primary source of AMR-related information at 55%. Media outlets, including television and social platforms, accounted for 30%. Community awareness initiatives represented 15%. Table 4 shows sources of AMR information.

**Table 4:** Sources of AMR Information.

| **Source** | **Frequency (%)** |
|---|---|
| Healthcare professionals | 55% |
| Media (TV, social media) | 30% |
| Community programs | 15% |

### 3.5 Socio-Demographic Influences on AMR Knowledge

Participants with advanced education exhibited enhanced AMR knowledge (p < .01). Higher-income individuals demonstrated increased adherence to complete antibiotic regimens (p < .05). Urban dwellers possessed superior AMR awareness compared to their rural counterparts (p < .05), while those with consistent healthcare access were less inclined to self-medicate (p < .01). Table 5 shows Socio-Demographic Influences on AMR Knowledge and Practices. **Figure 1 shows correlation matrix of Demographics and AMR Knowledge score.**

**Table 5:** Socio-Demographic Influences on AMR Knowledge and Practices.

| **Factor** | **Influence** | **p-value** |
|---|---|---|
| Education | Higher AMR knowledge | < .01 |
| Income | Greater adherence to prescriptions | < .05 |
| Urban vs. Rural | Higher AMR awareness in urban areas | < .05 |
| Healthcare access | Less self-medication with regular access | < .01 |

**Figure 1:**
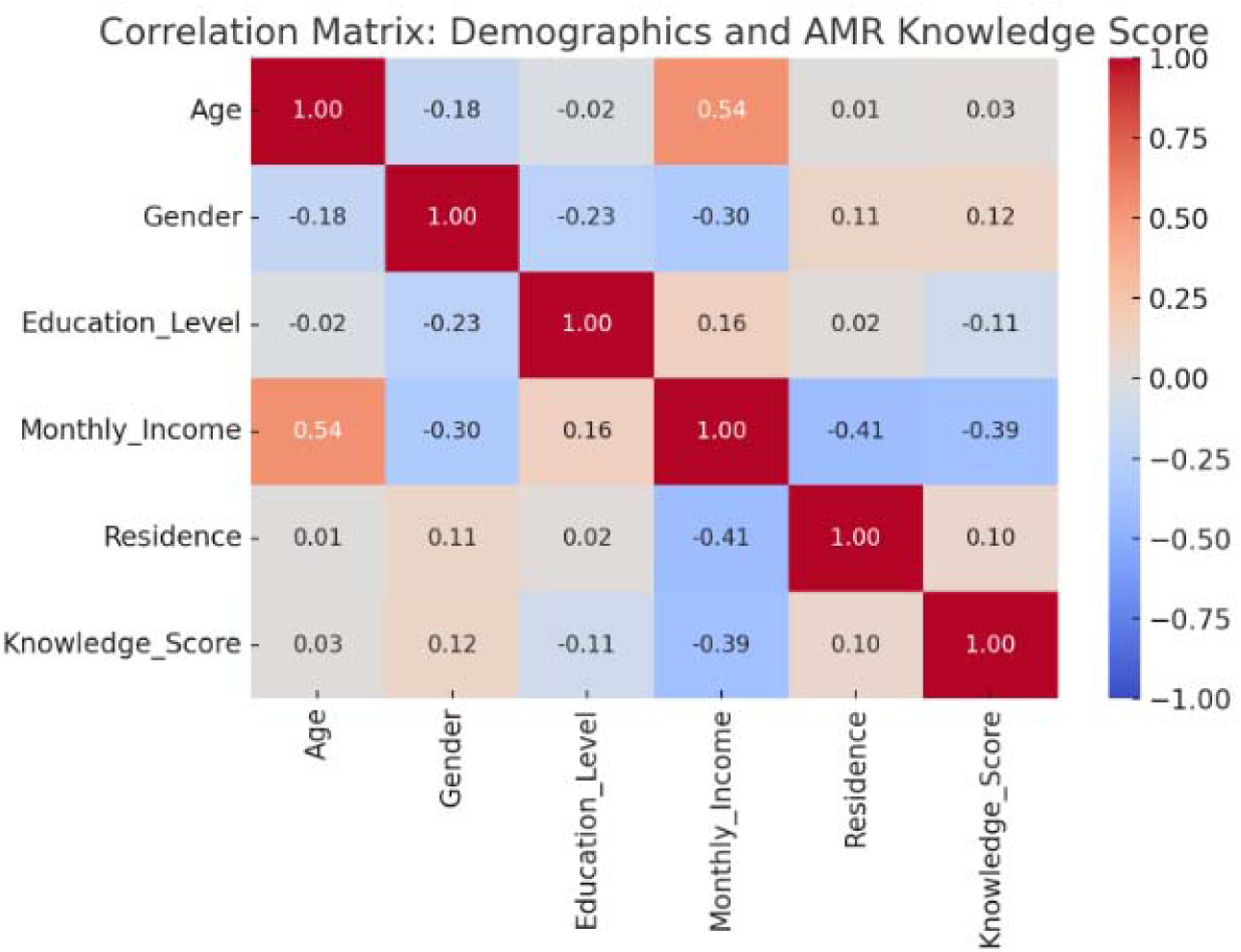
Demographics and AMR Knowledge score.

### 3.5 Chi-Square Test Findings

Chi-square analyses indicated significant links between education and proper antibiotic utilization (χ² = 12.4, p < .01), and between income level and adherence to prescriptions (χ² = 8.7, p < .05). Table 6 shows Chi-Square Test Results.

**Table 6:** Chi-Square Test Results.

| **Variable Pair** | **χ²** | **p-value** |
|---|---|---|
| Education × Antibiotic Use | 12.4 | < .01 |
| Income × Prescription Adherence | 8.7 | < .05 |

## 4. Discussion

This comprehensive investigation elucidates significant deficiencies in the knowledge and practices exhibited by caregivers with respect to the phenomenon of antimicrobial resistance (AMR) within the context of Bangladesh, a nation grappling with the complexities of public health challenges. Although there exists a relatively elevated level of awareness concerning the fundamental effectiveness of antibiotics in combating bacterial infections, it is concerning to note that substantial misconceptions endure, particularly with regard to the application of these medications for viral illnesses, as well as the premature discontinuation of prescribed treatment regimens before the full course has been completed. The findings of this study are consistent with those observed in other recent research endeavors conducted across South Asia, where the inappropriate utilization of antibiotics continues to represent a significant and pressing public health issue (Islam et al., 2023).

Over half of the participants in this study erroneously believed that the administration of antibiotics could expedite recovery from commonplace colds, or that it was acceptable to cease antibiotic treatment once the symptoms had subsided. Comparable misconceptions were identified in an extensive cross-sectional study involving parents in urban areas of Bangladesh, where a striking 60% of the participants were under the mistaken impression that antibiotics could shorten the duration of recovery from viral infections (Rahman et al., 2023). These entrenched beliefs substantially contribute to patterns of inappropriate antibiotic usage, which in turn drive the alarming development and dissemination of resistant bacterial strains that pose a significant threat to public health.

The investigation revealed that socio-demographic factors exert a notable influence on both the knowledge levels and behavioral patterns related to antibiotic usage among the participants. Individuals possessing higher levels of education and income were observed to display a superior understanding and adherence to established antibiotic guidelines when compared to their less advantaged counterparts. These findings corroborate existing evidence indicating that educational attainment and socio-economic status are critically influential in shaping health literacy and fostering responsible antibiotic behavior (Ahmed et al., 2024). Furthermore, urban respondents exhibited a higher level of awareness regarding AMR when juxtaposed with their rural counterparts, a disparity likely attributable to enhanced access to healthcare professionals and greater exposure to public awareness initiatives (Haque et al., 2022).

A disconcertingly high proportion—specifically 40%—of respondents acknowledged engaging in self-medication practices with antibiotics, frequently procuring these medications from pharmacies or utilizing leftover prescriptions from previous courses of treatment. This particular pattern of behavior mirrors national trends observed in Bangladesh, where antibiotics are commonly acquired without the necessity of a formal prescription owing to lax regulatory frameworks and the economic barriers that hinder access to formal healthcare services (World Health Organization [WHO], 2023). The over-the-counter availability of antibiotics presents a formidable challenge in low- and middle-income countries (LMICs), necessitating a concerted effort to address this issue through both stringent policy enforcement and comprehensive public education (Miller et al., 2024).

On a global scale, the issue of antimicrobial resistance is escalating at a disconcerting rate that warrants immediate attention and action. The Global Research on Antimicrobial Resistance (GRAM) Project has estimated that AMR was directly responsible for an alarming 1.27 million fatalities worldwide in the year 2019, with South Asia emerging as one of the most severely affected regions in this crisis (Murray et al., 2022). If the current trajectory of AMR continues unabated, it is projected that this phenomenon could result in as many as 10 million deaths annually by the year 2050, with profound implications for healthcare systems and economic stability globally (O’Neill, 2016). Therefore, the necessity for interventions in Bangladesh—a nation characterized by its densely populated and vulnerable demographics—carries substantial global significance that cannot be overlooked.

In order to effectively combat the burgeoning threat of AMR, it is imperative that a multifaceted approach is adopted, one that encompasses a variety of strategies and interventions. Specifically, targeted educational programs must be developed and implemented to address prevalent misconceptions surrounding antibiotic usage while simultaneously promoting responsible practices among caregivers. Additionally, it is critical to strengthen the regulatory framework governing antibiotic sales, enhance access to qualified healthcare providers, and increase investment in public health initiatives within rural areas, as these steps are essential for fostering a more informed and responsible approach to antibiotic utilization. As underscored by the World Health Organization (WHO, 2023), the One Health approach—which seeks to integrate the health of humans, animals, and the environment—is of paramount importance for achieving sustainable containment efforts against the pervasive threat of AMR.

## 5. Conclusion

This research highlights gaps concerning AMR knowledge and the inappropriate use of antibiotics by caretakers in Bangladesh. Most participants seemingly understood that antibiotics treat bacterial infections; however, they also practiced taking antibiotics for viral infections and discontinuing them before finishing the prescribed course. Respondents with more education, higher income, and living in urban areas exhibited better compliance with AMR and proper antibiotic practices.

Self-medication without adequate rationale remains the most common practice and increases the likelihood of AMR. Pre- and post-educational program evaluation data indicate that the rural and low literacy populations warmest attention make responsible and correct antibiotic use policies necessary. Milner also highlights the need to control the over-the-counter sale of antibiotics, reduced availability of primary health care services, and increased self-medication.

Addressing these issues is important for Bangladesh at the community level, particularly because of the risk AMR poses globally. There is a need for further research exploring how these initiatives increase long-term awareness and change behavior concerning AMR.

## Data Availability

All data produced in the present study are available upon reasonable request to the authors

## 6. Acknowledgement

We gratefully acknowledge the guidance and support of Mr. Faisal Ahmed, whose mentorship was crucial throughout the development and execution of this research project. We also extend our heartfelt thanks to the Bangladesh Institute of Innovative Health Research for their institutional support, resources, and ethical review that enabled this study to be conducted effectively. Finally, we appreciate the cooperation of all the participants and schools involved, without whom this research would not have been possible.

## 7. Funding

The authors received no financial support for the research, authorship, and/or publication of this article.

## Notes

### Competing Interest Statement

The authors have declared no competing interest.

### Author Declarations

The study was conducted in compliance with ethical standards and received approval from the Institutional Review Board (IRB) at the Bangladesh Institute of Innovative Health Research.

